# Self-reported smell and taste recovery in COVID-19 patients: a one-year prospective study

**DOI:** 10.1101/2021.03.18.21253862

**Authors:** Paolo Boscolo-Rizzo, Francesco Guida, Jerry Polesel, Alberto Vito Marcuzzo, Paolo Antonucci, Vincenzo Capriotti, Erica Sacchet, Fiordaliso Cragnolini, Andrea D’Alessandro, Enrico Zanelli, Riccardo Marzolino, Chiara Lazzarin, Margherita Tofanelli, Nicoletta Gardenal, Daniele Borsetto, Claire Hopkins, Luigi Angelo Vaira, Giancarlo Tirelli

## Abstract

**Purpose:** The aim of the present study was to estimate the one-year prevalence and recovery rate of self-reported chemosensory dysfunction in a series of subjects with previous mild-to-moderate symptomatic COVID-19.

**Methods:** Prospective study based on the SNOT-22 (item sense of smell or taste) and additional outcomes.

**Results:** 268/315 patients (85.1%) completing the survey at baseline also completed the follow-up interview. The 12-months prevalence of self-reported COVID-19 associated chemosensory dysfunction was 21.3% (95% CI: 16.5-26.7%). Of the 187 patients who complained of COVID-19 associated chemosensory dysfunction at baseline, 130 (69.5%; 95% CI 62.4-76.0%) reported complete resolution of smell or taste impairment, 41 (21.9%) reported a decrease in the severity, and 16 (8.6%) reported the symptom was unchanged or worse one year after onset. The risk of persistence was higher for patients reporting a baseline SNOT-22 score > o = 4 (OR=3.32; 95% CI: 1.32-8.36) as well as for those requiring > o = 22 days for a negative swab (OR=2.18; 95% CI: 1.12-4.27).

**Conclusion:** A substantial proportion of patients with previous mild-to-moderate symptomatic COVID-19 characterized by new onset of chemosensory dysfunction still complained on altered sense of smell or taste one-year after the onset.

## Introduction

One year after the outbreak of the coronavirus-19 disease (COVID-19) pandemic in Europe, alterations in smell and taste have unquestionably emerged as a highly prevalent symptom and the most reliable predictor of the severe acute respiratory syndrome–coronavirus-2 (SARS-CoV-2) infection [1–3]. In addition, a high persistent rate of chemosensory disorders has been reported 6 months after onset [4,5]. However, the results of these reports cannot be considered as definitive as in some forms of post-viral anosmia, recovery times of more than one year have been reported in the past [6].

Although psychophysical evaluation of the olfactory function has been observed to have higher sensitivity, especially in detecting mild hyposmia [4,7], the self-reported evaluation of chemosensitivity has a baseline parameter of comparison consisting in the subjective perception of smell or taste preceding the onset of COVID-19. In older adults, indeed, the prevalence of psychophysical olfactory impairment in the setting of no reported deficit is 15% [8].

The aim of the present study was to estimate the one-year prevalence and recovery rate of self-reported chemosensory dysfunction in a series of subjects with previous mild-to-moderate symptomatic COVID-19.

## Materials and Methods

The study was approved by the Ethics Committee of the Friuli Venezia Giulia Region (CEUR-2020-Os-156), and informed consent was obtained verbally for telephone interviews.

### Subjects

This is a prospective study on mild-to-moderate symptomatic adult patients consecutively assessed at Trieste University Hospital between March 1 and March 31, 2020, who tested positive for SARS-CoV-2 RNA by polymerase chain reaction (PCR) on nasopharyngeal and throat swabs performed according to World Health Organization recommendation [9]. All patients were initially home-isolated with mild-to-moderate symptoms. Patients were considered mildly-to-moderately symptomatic if they had less severe clinical symptoms with no evidence of pneumonia, not requiring hospitalization, and therefore considered suitable for being treated at home. Patients with a history of previous trauma, surgery or radiotherapy in the oral and nasal cavities, allergic rhinitis or rhinosinusitis, previous olfactory or gustative dysfunction, or psychiatric or neurological diseases, were excluded from the study.

315 (72.7%) of the 433 eligible patients completed the baseline telephone interview administrated within 3 weeks after the first positive swab performed between 1 – 22 March 2021. The median time from symptoms onset to SARS-CoV-2 testing was 7 days (interquartile range, 4-11 days). All patients completing the baseline interview were phoned from 5th to 12th March 2021, so that all patients were recontacted 12 months after the onset of symptoms; in case of a non-response, patients were re-contacted twice.

### Questionnaires

Demographic and clinical data were collected through ad hoc questions administered during the baseline interview and included gender, age, self-reported height and weight, smoking and alcohol habits, and the following co-morbidities: immunosuppression, diabetes, cardiovascular diseases, active cancer, chronic respiratory disease, kidney disease, liver disease. Obesity was defined as having a body mass index (BMI) of 30 or more. The sense of smell and taste was assessed by the Sino-Nasal Outcome test 22 (SNOT-22)[10], item “sense of smell or taste”, both at baseline and during the follow-up interview to evaluate their persistence and the recovery rate. The SNOT-22 grades symptom severity as none (0), very mild (1), mild or slight (2), moderate (3), severe (4), or as bad as it can be (5). Patients with SNOT-22 > 0 were also asked whether the chemosensory alteration involved the sense of smell, taste, or both. Patients were finally asked about blocked nose based on the item scores No= 0, Yes—mild to moderate = 1, and Yes—severe = 2. The dates of the first positive and negative swabs were obtained from hospital records.

### Statistical analysis

Symptom prevalence was expressed as percentage of total patients, and 95% confidence interval (CI) were calculated using Clopper-Pearson method; differences in prevalence were evaluated through Fisher’s exact test. The risk of chemosensory impairment persistence, expressed as odds ratio (OR), was estimated through unconditional logistic regression model, adjusting for sex and age. Variables which were significant ant the univariate analysis were further included in the multivariable model. Analyses were performed using R 3.6. and statistical significance was claimed for p<0.05 (two-tailed).

## Results

Of 315 patients completing the survey at baseline, 47 did not answer or refused the follow-up interview, thus leaving 268 responders (85.1%). Baseline socio-demographic and clinical characteristics of 268 patients are reported in Table 1. The median age of the study cohort was 48 years (interquartile range, 38-56 years). There was a female preponderance with 166 out of 268 being females (61.9%). Associated co-morbidities were reported by 91 cases (34.0%) with the most common being obesity reported by 32 patients (11.9%) followed by cardiovascular diseases (n=25, 9.3%) and immunosuppression (n=19, 7.1%).

**Table 1.** Baseline sociodemographic and clinical characteristics of 268 patients positive for SARS-CoV-2.

|  | n | % | (95% CI) |
| --- | --- | --- | --- |
| Gender |  |  |  |
| Male | 102 | 38.1 | (32.2-44.2) |
| Female | 166 | 61.9 | (55.8-67.8) |
| Age (years) |  |  |  |
| <40 | 79 | 29.5 | (24.1-35.3) |
| 40-54 | 103 | 38.4 | (32.6-44.5) |
| ≥55 | 86 | 32.1 | (26.5-38.0) |
| Smoking habits |  |  |  |
| Never | 157 | 58.6 | (52.4-64.5) |
| Former | 68 | 25.4 | (20.3-31.0) |
| Current | 43 | 16.0 | (11.9-21.0) |
| Drinking habits |  |  |  |
| Never | 176 | 65.7 | (59.7-71.3) |
| Former | 27 | 10.1 | (6.7-14.3) |
| Current | 65 | 24.3 | (19.2-29.8) |
| BMI (kg m<sup>-2</sup>) |  |  |  |
| <25 | 146 | 54.5 | (48.3-60.5) |
| 25-29.9 | 84 | 31.3 | (25.8-37.3) |
| ≥30 | 38 | 14.2 | (10.2-18.9) |
| Comorbidity |  |  |  |
| None | 177 | 66.0 | (60.0-76.0) |
| Any | 91 | 34.0 | (28.3-40.0) |
| Blocked nose |  |  |  |
| 0: No | 218 | 81.3 | (76.2-85.8) |
| 1: Yes-mild to moderate | 35 | 13.1 | (9.3-17.7) |
| 2: Yes-severe | 15 | 5.6 | (3.2-9.1) |
| Chemosensory dysfunction |  |  |  |
| No | 81 | 30.2 | (24.8-36.1) |
| Yes | 187 | 69.8 | (63.9-75.2) |
| Type of chemosensory dysfunction |  |  |  |
| None | 81 | 30.2 | (24.8-36.1) |
| Smell | 19 | 7.1 | (4.3-10.9) |
| Taste | 16 | 6.0 | (3.5-9.5) |
| Smell and taste | 152 | 56.7 | (50.6-62.7) |
| SNOT-22 at diagnosis<sup>a</sup> |  |  |  |
| 0: No | 81 | 30.2 | (24.8-36.1) |
| 1: Very mild | 3 | 1.1 | (0.2-3.2) |
| 2: Mild or slight | 18 | 6.7 | (4.0-10.4) |
| 3: Moderate | 27 | 10.1 | (6.7-14.3) |
| 4: Severe | 40 | 14.9 | (10.9-19.8) |
| 5: As bad as it can be | 99 | 36.9 | (31.1-43.0) |
| Time for negative swab (days) |  |  |  |
| <22 | 128 | 47.8 | (41.6-53.9) |
| ≥22 | 140 | 52.2 | (46.1-58.4) |

Abbreviation: SARS-CoV-2, severe acute respiratory syndrome coronavirus
<sup>a</sup>According to Sino-Nasal Outcome Test 22 (SNOT-22), item “sense of smell or taste”

The median time to achieving a negative swab was 22 days (interquartile range, 15-31 days). Overall, 187 of 268 responders (69.8%, CI 95% 63.9-75.2) reported an altered sense of smell or taste at baseline (SNOT-22 > 0) with 99 (36.9%, CI 95% 31.1-43.0) reporting the highest SNOT-22 score, for severe problem. Particularly, 152 subjects (81.3%) self-reported combined chemosensory dysfunction, 19 (10.2%) patients self-reported isolated smell impairment and 16 patients (8.6%) self-reported isolated taste disorder. No sociodemographic characteristic or clinical feature was associated with either chemosensory dysfunctions or its severity at baseline (data not shown).

After 12 months, 57 patients (21.3%; 95% CI: 16.5-26.7%) still reported chemosensory dysfunction, with 34 subjects still reporting both smell and taste dysfunction, 15 reporting smell impairment and 5 taste disorder. Among patients with persistent chemosensory dysfunction, only 4 (7.0%) complained blocked nose. Among the 187 patients who have complained a COVID-19 associated chemosensory dysfunction at baseline, 130 (69.5%; 95% CI 62.4-76.0%) reported complete resolution of smell or taste impairment, 41 (21.9%; 95% CI: 16.2-28.5%) reported a decrease in the severity, and 16 (8.6%; 95% CI: 5.0-13.5%) reported the symptom was unchanged or worse (Table 2). Baseline socio-demographic and lifestyle factors were not associated with the persistence of chemosensory dysfunction (Table 3). After adjustment for covariates, the severity of chemosensory dysfunction at baseline (OR=3.32; 95% CI: 1.32-8.36 for SNOT-22 score ≥ 4) and a longer time to achieving a negative swab (OR=2.18; 95% CI: 1.12-4.27) were associated with a higher risk of persistence of symptoms at 12-months.

**Table 2.**
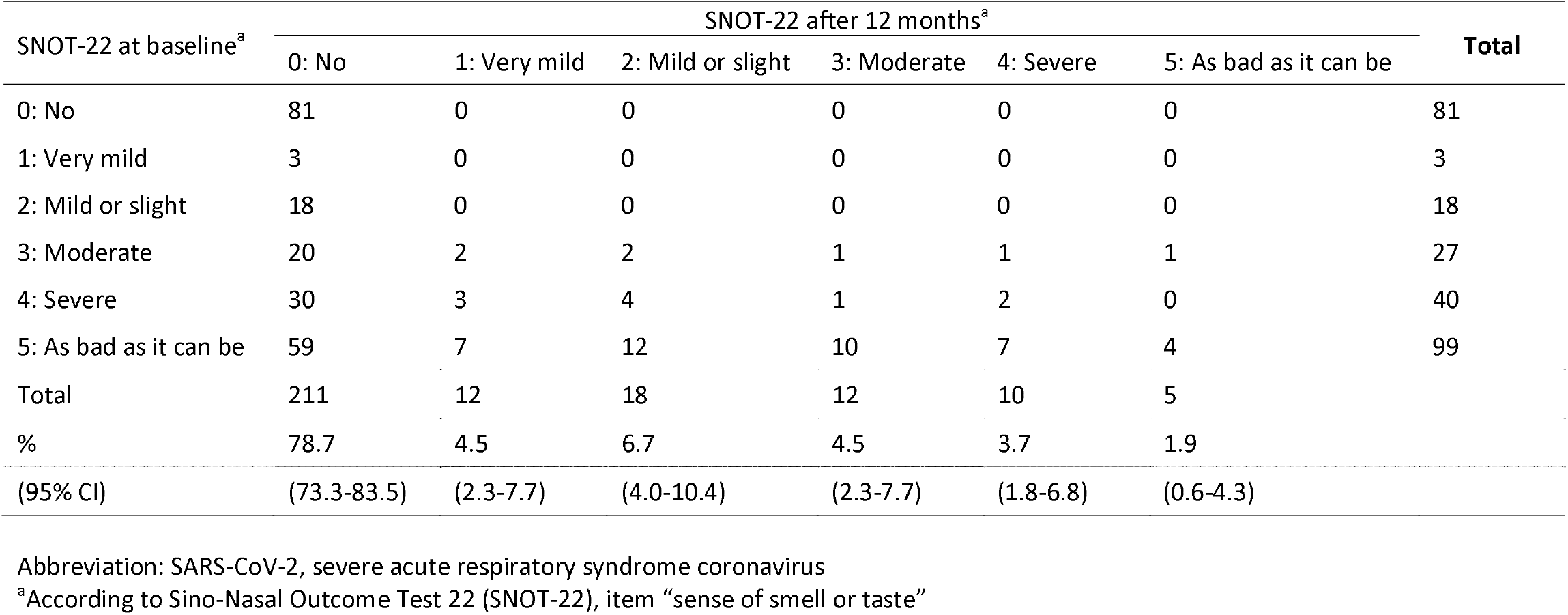
Change in alteration of sense of smell or taste in 268 patients positive for SARS-CoV-2.

| SNOT-22 at baseline<sup>a</sup> | SNOT-22 after 12 months<sup>a</sup> |  |  |  |  |  | Total |
| --- | --- | --- | --- | --- | --- | --- | --- |
|  | 0: No | 1: Very mild | 2: Mild or slight | 3: Moderate | 4: Severe | 5: As bad as it can be |  |
| 0: No | 81 | 0 | 0 | 0 | 0 | 0 | 81 |
| 1: Very mild | 3 | 0 | 0 | 0 | 0 | 0 | 3 |
| 2: Mild or slight | 18 | 0 | 0 | 0 | 0 | 0 | 18 |
| 3: Moderate | 20 | 2 | 2 | 1 | 1 | 1 | 27 |
| 4: Severe | 30 | 3 | 4 | 1 | 2 | 0 | 40 |
| 5: As bad as it can be | 59 | 7 | 12 | 10 | 7 | 4 | 99 |
| Total | 211 | 12 | 18 | 12 | 10 | 5 |  |
| % | 78.7 | 4.5 | 6.7 | 4.5 | 3.7 | 1.9 |  |
| (95% CI) | (73.3-83.5) | (2.3-7.7) | (4.0-10.4) | (2.3-7.7) | (1.8-6.8) | (0.6-4.3) |  |

Abbreviation: SARS-CoV-2, severe acute respiratory syndrome coronavirus
<sup>a</sup>According to Sino-Nasal Outcome Test 22 (SNOT-22), item “sense of smell or taste”

**Table 3.** Odds ratio (OR) and 95% confidence interval (CI) for persistence/worsening at 12 month of alteration of sense of smell or taste according to baseline characteristics

| Characteristics | Persistence/<br>worsening |  | Recovered |  | OR (95% CI)<sup>a</sup> | OR (95% CI)<sup>b</sup> |
| --- | --- | --- | --- | --- | --- | --- |
|  | n | (%) | n | (%) |  |  |
| Gender |  |  |  |  |  |  |
| Male | 17 | (29.8) | 53 | (40.8) | Reference | Reference |
| Female | 40 | (70.2) | 77 | (59.2) | 1.67 (0.85-3.27) | 1.83 (0.90-3.72) |
| Age (years) |  |  |  |  |  |  |
| <40 | 20 | (35.1) | 35 | (26.9) | Reference | Reference |
| 40-54 | 21 | (36.8) | 55 | (42.3) | 0.64 (0.30-1.35) | 0.49 (0.22-1.11) |
| ≥55 | 19 | (28.1) | 40 | (30.8) | 0.69 (0.31-1.54) | 0.59 (0.25-1.38) |
| Smoking habits |  |  |  |  |  |  |
| Never | 30 | (52.6) | 84 | (64.6) | Reference | Reference |
| Ever | 27 | (47.4) | 46 | (35.4) | 1.77 (0.93-3.38) | 1.77 (0.89-3.51) |
| Drinking habits |  |  |  |  |  |  |
| Never | 42 | (73.7) | 79 | (60.8) | Reference | Reference |
| Ever | 15 | (26.3) | 41 | (39.2) | 0.60 (0.29-1.21) | 0.67 (0.32-1.42) |
| BMI (kg m<sup>-2</sup>) |  |  |  |  |  |  |
| <30 | 35 | (61.4) | 59 | (45.4) | Reference | Reference |
| ≥30 | 22 | (38.6) | 71 | (54.6) | 0.72 (0.30-1.75) | 0.55 (0.22-1.40) |
| Blocked nose |  |  |  |  |  |  |
| 0 | 40 | (70.2) | 107 | (82.3) | Reference | Reference |
| 1-2 | 17 | (29.8) | 23 | (17.7) | 1.97 (0.95-4.11) | 2.14 (0.98-4.67) |
| Comorbidity |  |  |  |  |  |  |
| None | 39 | (68.4) | 81 | (62.3) | Reference | Reference |
| Any | 18 | (31.6) | 49 | (37.7) | 0.80 (0.41-1.59) | 0.84 (0.40-1.74) |
| Type of chemosensory impairment |  |  |  |  |  |  |
| Smell or taste | 5 | (8.8) | 30 | (23.1) | Reference | Reference |
| Smell and taste | 52 | (91.2) | 100 | (76.9) | 3.37 (1.22-9.33) | 2.58 (0.91-7.37) |
| SNOT-22<sup>b</sup> |  |  |  |  |  |  |
| 1-3 | 7 | (12.3) | 41 | (31.5) | Reference | Reference |
| 4-5 | 50 | (87.7) | 89 | (68.5) | 3.84 (1.55-9.49) | 3.32 (1.32-8.36) |
| Time for negative swab (days) |  |  |  |  |  |  |
| <22 | 21 | (36.8) | 70 | (53.9) | Reference | Reference |
| ≥22 | 36 | (63.2) | 60 | (46.2) | 2.09 (1.09-3.99) | 2.18 (1.12-4.27) |

Abbreviation: BMI, body mass index
<sup>a</sup>Estimated from unconditional logistic regression model, adjusting for sex and age. <sup>b</sup>Further adjusted for smell/taste alteration, severity of alteration, and time for negative swab
<sup>b</sup>According to Sino-Nasal Outcome Test 22 (SNOT-22), item “sense of smell or taste”

## Discussion

Among the 187 patients who have complained a COVID-19 associated chemosensory dysfunction, 69.5% experienced a complete resolution of these symptoms at 12-months while 30.5% still reported impairment in their chemosensory function. Thus, the 12-months overall prevalence of persistent altered sense of smell or taste in this series of subjects with previous mild-to-moderate symptomatic COVID-19 was 21.3%.

To the best of our knowledge this is the first study to estimate the 12-months prevalence and recovery rate of self-reported chemosensory dysfunction in a series of subjects with previous mild-to-moderate symptomatic SARS-CoV-2 infection. Such a long follow-up is essential to estimate the real prevalence of COVID-19 related chemosensory disorders as functional recoveries after one year from infection with other viruses have been reported in the past even [6].

The data from our study highlights a significant rate of lasting olfactory dysfunction as a legacy of the pandemic. Considering the spread out of SARS-CoV-2 infections in Europe with more than 36,000,000 cases to date [11], the burden of chemosensory disorders on the health systems will be even more pressing. Indeed, given that our estimates are based on self-reported symptoms, these patients will likely seek medical care for their chemosensory disorders.

This prevalence is likely underestimated compared to the rate that would have been obtained using the most accurate psychophysical evaluation [4,12]. However, this subjective evaluation may be more accurate in estimating the percentage of patients in whom the smell or taste disturbance has repercussions on their quality of life. Furthermore, the evaluation with psychophysical tests alone could overestimate the prevalence of residual COVID-19 related chemosensory disorders including all those subjects who were unaware of having a previous olfactory or gustatory dysfunction [8].

These observations should prompt the experts in chemosensory disorders to make further efforts to test possible treatments for post-COVID-19 smell and taste disorders. Current therapies are essentially based on the experience gained in the study of other forms of post-viral anosmia. Recently, the Clinical Olfactory Working Group members made an overwhelming recommendation for olfactory training for post-viral anosmia including COVID-19 [13]. However, it is imperative to design multicentre clinical trials to test olfactory training efficacy as well as new therapeutic strategies for improving chemosensory outcomes in patients with post-COVID-19 chemosensory disorders. The high number of potentially recruitable patients offers a unique opportunity to test the efficacy of new therapeutic approaches for post-viral chemosensory disorders.

We found that the severity of chemosensory dysfunction at baseline was associated with a higher risk of persistence of symptoms at 12-months. Thus, patients with more severe smell or taste impairment should be preferentially included in randomized clinical trials evaluation the efficacy of different treatment approaches, and research is required to determine of treatment at an early stage can reduce the rate of persistent dysfunction.

We also found that a longer duration of viral persistence on PCR testing was associated with an increased risk of persistent chemosensory dysfunction. It has been shown that SARS-CoV-2 gains entry to the supporting ells of the respiratory epithelium [14]. It could be hypothesized that persistence of viral infection in the nose prevents recovery of the supporting cells, which leads indirectly to injury of the olfactory sensory neurones and persistent loss. In contrast, rapid viral clearance may allow repair of the integrity of the olfactory epithelium before loss of the olfactory sensory neurons occurs.

The data form the present study should be taken cautiously owing to several study limitations. Symptoms were self-reported, based on cross-sectional surveys, and may therefore contain suboptimal sensitivity. Subjective evaluation of the olfactory function was observed, indeed, to be inadequate to fully evaluate olfactory recovery with several authors having underlined that subjectivity of self-reporting may lead to underestimation of the prevalence of olfactory dysfunction. It is clear that patients’ rating of whether smell and or taste are impacted differentially is very difficult to interpret, due to impact of loss perception of flavour through retronasal olfaction, and little can be inferred from this aspect of the study. Moreover, the study cohort was relatively small and patients with more severe COVID-19 were excluded.

In conclusion, a substantial proportion of patients with previous mild-to-moderate symptomatic COVID-19 characterized by new onset of chemosensory dysfunction still complained on altered sense of smell or taste one-year after the onset. As recovery from post-viral loss may continue beyond this period [6,15], further assessments will be necessary to conclude whether smell and taste dysfunction is permanent. Moreover, there is urgent need for more efforts and research on treatment strategies for post-COVID-19 chemosensory dysfunction.

## Data Availability

The authors confirm that the data supporting the findings of this study are available within the article. Raw data are available upon request.

## References

[1] Lechien JR, Chiesa-Estomba CM, De Siati DR, Horoi M, Le Bon SD, Rodriguez A, et al. Olfactory and gustatory dysfunctions as a clinical presentation of mild-to-moderate forms of the coronavirus disease (COVID-19): a multicenter European study. Eur Arch Oto-Rhino-Laryngol Off J Eur Fed Oto-Rhino-Laryngol Soc EUFOS Affil Ger Soc Oto-Rhino-Laryngol - Head Neck Surg 2020. https://doi.org/10.1007/s00405-020-05965-1.

[2] Spinato G, Fabbris C, Polesel J, Cazzador D, Borsetto D, Hopkins C, et al. Alterations in Smell or Taste in Mildly Symptomatic Outpatients With SARS-CoV-2 Infection. JAMA 2020. https://doi.org/10.1001/jama.2020.6771.

[3] Gerkin RC, Ohla K, Veldhuizen MG, Joseph PV, Kelly CE, Bakke AJ, et al. Recent smell loss is the best predictor of COVID-19 among individuals with recent respiratory symptoms. Chem Senses 2020. https://doi.org/10.1093/chemse/bjaa081.

[4] Boscolo-Rizzo P, Menegaldo A, Fabbris C, Spinato G, Borsetto D, Vaira LA, et al. Six-month psychophysical evaluation of olfactory dysfunction in patients with COVID-19. Chem Senses 2021. https://doi.org/10.1093/chemse/bjab006.

[5] Hopkins C, Surda P, Vaira LA, Lechien JR, Safarian M, Saussez S, et al. Six month follow-up of self-reported loss of smell during the COVID-19 pandemic. Rhinology 2020. https://doi.org/10.4193/Rhin20.544.

[6] Lee DY, Lee WH, Wee JH, Kim J-W. Prognosis of postviral olfactory loss: follow-up study for longer than one year. Am J Rhinol Allergy 201428:419–22. https://doi.org/10.2500/ajra.2014.28.4102.

[7] Vaira LA, Deiana G, Fois AG, Pirina P, Madeddu G, De Vito A, et al. Objective evaluation of anosmia and ageusia in COVID-19 patients: Single-center experience on 72 cases. Head Neck 2020. https://doi.org/10.1002/hed.26204.

[8] Murphy C, Schubert CR, Cruickshanks KJ, Klein BEK, Klein R, Nondahl DM. Prevalence of olfactory impairment in older adults. JAMA 2002288:2307–12. https://doi.org/10.1001/jama.288.18.2307.

[9] Technical guidance n.d. https://www.who.int/emergencies/diseases/novel-coronavirus-2019/technical-guidance (accessed April 27, 2020).

[10] Hopkins C, Gillett S, Slack R, Lund VJ, Browne JP. Psychometric validity of the 22-item Sinonasal Outcome Test. Clin Otolaryngol Off J ENT-UK Off J Neth Soc Oto-Rhino-Laryngol Cervico-Facial Surg 200934:447–54. https://doi.org/10.1111/j.1749-4486.2009.01995.x.

[11] Coronavirus Update (Live): 120,126,411 Cases and 2,661,042 Deaths from COVID-19 Virus Pandemic - Worldometer n.d. https://www.worldometers.info/coronavirus/ (accessed March 14, 2021).

[12] Vaira LA, Hopkins C, Petrocelli M, Lechien JR, Chiesa-Estomba CM, Salzano G, et al. Smell and taste recovery in coronavirus disease 2019 patients: a 60-day objective and prospective study. J Laryngol Otol 2020134:703–9. https://doi.org/10.1017/S0022215120001826.

[13] Addison AB, Wong B, Ahmed T, Macchi A, Konstantinidis I, Huart C, et al. Clinical Olfactory Working Group consensus statement on the treatment of postinfectious olfactory dysfunction. J Allergy Clin Immunol 2021. https://doi.org/10.1016/j.jaci.2020.12.641.

[14] Brann DH, Tsukahara T, Weinreb C, Lipovsek M, Van den Berge K, Gong B, et al. Non-neuronal expression of SARS-CoV-2 entry genes in the olfactory system suggests mechanisms underlying COVID-19-associated anosmia. Sci Adv 2020;6. https://doi.org/10.1126/sciadv.abc5801.

[15] Reden J, Mueller A, Mueller C, Konstantinidis I, Frasnelli J, Landis BN, et al. Recovery of olfactory function following closed head injury or infections of the upper respiratory tract. Arch Otolaryngol Head Neck Surg 2006;132:265–9. https://doi.org/10.1001/archotol.132.3.265.

